# Utilizing artificial intelligence-driven virtual standardized pediatric patients to enhance the capabilities of primary healthcare doctors in China for managing common pediatric diseases: a study protocol for a randomized controlled trial

**DOI:** 10.1101/2024.10.31.24315838

**Authors:** Yanna Mao, Huanyuan Luo, Yiyuan Cai, Wanqing Huang, Fang Fang, Yue Lu, Xin Chen, Qing Zhao, Duolao Wang, Hua He, Xiaohui Wang, Dexing Zhang, Guobao Li, Yichi Zhang, Dong (Roman) Xu, Yao Zhao

## Abstract

**Background:** China’s healthcare system for children faces significant challenges, particularly due to the limited pediatric service capacity of primary healthcare institutions. A shortage of effective and accessible training tools for primary care doctors further hinders progress in addressing this gap. Technological advancements, especially in artificial intelligence, offer a potential solution to improve pediatric care. Artificial intelligence-driven virtual standardized patients (VSPs), leveraging internet and virtual simulation technologies, simulate clinical cases with specific disease characteristics, providing an innovative, efficient, and flexible training method. VSPs are increasingly utilized in medical education, clinical reasoning, and licensure exams. This study focuses on using VSPs to improve the management of common pediatric conditions, which are major health concerns for children and impose significant psychological and financial burdens on families.

**Methods:** This study will involve a three-arm randomized controlled trial to evaluate the effectiveness of a virtual pediatric standardized patient platform in enhancing primary care doctors’ management of common pediatric diseases. At least 459 participants, including general practitioners, internal medicine practitioners, surgeons, and pediatricians from more than 10 provinces across China, will be randomly assigned to one of three groups: the virtual patient platform group, the case teaching manual group, or the case teaching video group. Five virtual patient cases covering pneumococcal pneumonia, rotavirus enteritis with hypovolemic shock, hand-foot-and-mouth disease, acute appendicitis, and respiratory failure will be developed, along with corresponding case teaching materials. After a two-week learning period, participants’ disease management abilities will be assessed using clinical vignettes. The primary outcome is adherence to best clinical practice guidelines, categorized into full adherence, partial adherence, and nonadherence.

**Discussion:** This study aims to leverage artificial intelligence for capacity enhancement, targeting the shortcomings of primary care pediatrics and using VSP to help enhance primary care pediatrics capacity. It is a randomized controlled trial involving over 300 primary healthcare institutions across more than 10 provinces in China, ensuring broad and representative participation from both developed and underdeveloped regions.

**Trial registration:** Chictr.org.cn, <u>ChiCTR2400085808</u>. Registered on 19 June 2024.

## 1. Background

Currently, China’s healthcare system for children is not fully developed. One of the primary obstacles is the diminished capacity of primary healthcare institutions to provide pediatric services [1]. The pediatric capabilities of primary care doctors in China are insufficient, and there is a lack of convenient and effective training tools, which hinders the implementation of the “Healthy China” strategy. However, with technological advancements, artificial intelligence can efficiently and cost-effectively enhance the pediatric service capabilities of primary care doctors.

Virtual standardized patients (VSPs), utilizing internet and virtual simulation technology, emulate patients with specific disease characteristics and clinical manifestations. They represent an advanced interdisciplinary field that merges digital technology with medical science. By leveraging these technologies, VSPs facilitate simulations of entities with disease characteristics, marking a revolutionary development in biomedicine. VSPs models can expand subject cohorts, accelerate development, improve efficiency, provide evidence for the safety and efficacy of treatment plans, support precision medicine, and enable cost-effective planning [2]. With advantages in safety, flexibility, convenience, and efficiency, VSPs are widely used in medical education, clinical reasoning training, and licensure examinations. In this project, doctors will interact with VSPs to conduct clinical simulations and training, including consultations, physical examinations, auxiliary examinations, and treatment decision-making.

Common pediatric diseases include pneumococcal pneumonia, rotavirus enteritis with hypovolemic shock, hand-foot-and-mouth disease (HFMD), acute appendicitis, and respiratory failure. These conditions pose significant health risks to children and create psychological anxiety and financial burdens for their families.

Streptococcus pneumoniae is a major pathogen responsible for respiratory tract infections and invasive diseases in the community. It can lead to various conditions such as otitis media, sinusitis, pneumonia, bacteremia, and meningitis. Children and the elderly are particularly susceptible to pneumococcal infections. In China, Streptococcus pneumoniae is the most common pathogen causing community-acquired pneumonia in children aged 20 days to school age, as well as severe pneumonia and necrotizing pneumonia [3, 4]. Evaluations of cause-specific mortality in children have shown that the case fatality rate for pneumonia in China can reach 30.5% [5].

Diarrhea caused by rotavirus infection, also known as rotavirus enteritis, can be life-threatening due to hypovolemic shock if left untreated. Diarrhea is a gastrointestinal syndrome characterized by increased stool frequency and changes in stool consistency, and is one of the most common diseases among infants and young children in China. Common complications include malnutrition, anemia, and electrolyte disturbances [6].

HFMD is a contagious viral infection in children, characterized by rashes and blisters on the hands, feet, mouth, and buttocks, often accompanied by fever [7]. The disease is highly infectious and spreads rapidly, particularly during peak seasons. A small percentage of severe cases may lead to serious complications, such as myocarditis, which require timely treatment. An investigation by Liu et al. reported a total of 442,079 cases of HFMD in Sichuan Province from 2017 to 2021, with an incidence rate of 106 per 100,000 people [^8^]. Among these cases, there were 627 severe cases and 4 deaths, with children under five years old accounting for 90% of the cases. A study on the economic burden and impact of HFMD in Bao’an District, Shenzhen, involving 1,225 patients, found that the overall economic burden for 1,195 outpatient cases was 422 RMB per case, while for 30 hospitalized cases it was (4386±1624) RMB per case. The study indicated that the economic burden for hospitalized patients was significantly higher than that for outpatients, with factors such as age, type of reimbursement, and number of visits influencing the economic burden of outpatient HFMD cases [9]. Early detection, diagnosis, and treatment can effectively reduce the economic burden of the disease.

Acute appendicitis is an acute abdominal condition that can arise from multiple factors, commonly occurring in children. It often presents with atypical clinical symptoms and progresses rapidly. If not treated promptly, it can lead to serious complications, including sepsis and septic shock, with some severe cases resulting in death [10]. Acute appendicitis is a common pediatric surgical emergency, accounting for 1% to 8% of all cases of abdominal pain [11]. When abdominal inflammation occurs in children, it frequently progresses to perforation or peritonitis due to delayed diagnosis [12].

Respiratory failure is a heterogeneous clinical syndrome associated with high mortality and long-term disability rates. It is one of the common severe conditions encountered in pediatrics. Acute respiratory distress syndrome, a specific type of pediatric respiratory failure, is frequently observed in pediatric intensive care units and poses a serious threat to children’s health. Therefore, standardized diagnosis and management are essential. While the clinical manifestations of respiratory failure may vary depending on the underlying disease, they prominently include changes in respiration, along with two major syndromes of hypoxemia and hypercapnia [13]. In emergency situations, children with impaired respiratory function may require immediate assessment and stabilization.

This study aims to develop a virtual standardized pediatric patient platform and conduct a randomized controlled trial comparing it with traditional case teaching manuals and videos. The research will explore the effectiveness of the virtual standardized pediatric patient platform in enhancing the ability of primary care doctors in China to manage common pediatric diseases, including pneumococcal pneumonia, rotavirus enteritis with hypovolemic shock, HFMD, acute appendicitis, and respiratory failure. The findings will provide scientific evidence for the education and training of primary care doctors and offer guidance and support to improve pediatric service levels in primary care settings.

## 2. Research design and methods

### 2.1. Research Design

This study proposes to conduct a three-arm randomized controlled trial. Participants will be randomly assigned to one of three groups: the virtual standardized pediatric patient group, the case teaching manual group, and the case teaching video group. Following a two-week learning period, the pediatric disease management abilities of doctors in each group will be evaluated.

### 2.2. Research Objectives

The primary objective of this study is to validate the effectiveness of a virtual standardized pediatric patient platform in enhancing primary care doctors’ abilities to manage common pediatric diseases. The research team will develop five virtual standardized patient cases, focusing on pneumococcal pneumonia, rotavirus enteritis with hypovolemic shock, hand-foot-and-mouth disease, acute appendicitis, and respiratory failure. In conjunction with these cases, corresponding case teaching manuals and videos will be created, establishing three training groups for primary care doctors. Subsequently, the research team will conduct a randomized controlled trial to evaluate the effectiveness of the virtual pediatric patient platform in enhancing primary care doctors’ management skills for common pediatric conditions, compared to traditional training methods of case teaching manuals and videos.

### 2.3. Research subjects

The main research subjects of this study are general practitioners, internal medicine practitioners, surgeons, and pediatricians working in primary medical institutions.

#### 2.3.1. Inclusion and Exclusion Criteria for Research Institutions

Inclusion Criteria: Primary and secondary hospitals, community health centers (stations) and clinics, as well as township health centers and village health clinics.

Exclusion Criteria: Specialized medical institutions (such as specialized hospitals and dental clinics), public health prevention and treatment institutions (such as tuberculosis prevention centers), ethnic medicine institutions (such as Mongolian and Tibetan hospitals), hospitals above the secondary level, and hospitals that have not yet been graded (due to their short establishment time and potentially unstable operations).

#### 2.3.2. Inclusion and exclusion criteria for research subjects

Inclusion criteria: Practicing (assistant) doctors and rural doctors working in medical institutions that meet the above conditions, with a scope of practice only including general practice, internal medicine, surgery, and pediatrics.

Exclusion criteria: None.

### 2.4. Interventions

Intervention group: Virtual standardized pediatric patients (VSPs). This group will engage with virtual standardized patient cases representing five conditions: pneumococcal pneumonia, rotavirus enteritis with hypovolemic shock, hand-foot-and-mouth disease, acute appendicitis, and respiratory failure. Primary care doctors in the intervention group will interact with virtual standardized patients to conduct interviews, physical examinations, auxiliary examinations, diagnoses, and treatments, simulating real clinical scenarios.

Control group 1: Case teaching manuals. The research team has developed knowledge manuals based on the same five conditions. The teaching objectives and content of these manuals align with those of the virtual standardized pediatric patients.

Control group 2: Case teaching videos. The research team has created teaching videos based on the same five conditions. The teaching objectives and content of these videos are consistent with those of the virtual pediatric patients.

### 2.5. Sample Size Calculation

Sample size calculation is based on the primary outcome, doctor adherence to best clinical practice guidelines, i.e., the extent to which doctors consistently make judgments and treatments based on best clinical practice guidelines and progression of the disease (an ordered categorical variable, consisting of three grades: full adherence, partial adherence, and nonadherence), which is measured using clinical vignette. Previously, we conducted a pilot study based on 29 doctors or pediatric medical postgraduates and obtained the results of the primary outcome for the intervention group and two control groups (Table 1). The pilot study showed that doctors and medical postgraduates were 10.46 times more likely to increase their adherence to best clinical practice guidelines by one level through VSP training than through video training and 17.98 times more likely to increase their adherence than through textbook training (Table 2). The test power is set at 0.8. This study will include a multiple group comparison among the three training groups, and a Bonferroni correction is used to minimize the risk of Type I errors, so the corrected test level is set at α=0.05/2=0.025 (two-sided). The sample size is calculated using the “Tests for Two Ordered Categorical Variables” command in the PASS software, and taking into account the possibility of subject attrition and other factors that may lead to a dropout rate of approximately 35%, the sample size required to compare the VSP with the video group is 153 participants per group, and the sample size required to compare the VSP with the manual group is 71 participants per group. So it is decided to set the sample size at 153 participants per group, resulting in an estimated total sample size of 459 participants for the three groups.

**Table 1:** Results of primary outcome in pilot study.

|  | VSP training | Video training | Manual training | All |
| --- | --- | --- | --- | --- |
| N | 10 | 9 | 10 | 29 |
| Nonadherence, n/(%) | 1(10.0) | 1(11.1) | 4(40.0) | 6(20.7) |
| Partial adherence, n/(%) | 2(20.0) | 2(22.2) | 2(20.0) | 6(20.7) |
| Full adherence, n/(%) | 7(70.0) | 6(66.7) | 4(40.0) | 17(58.6) |

**Table 2:** Results of sample size calculation.

|  | Odds ratio<br>estimated in<br>pilot study | Sample size<br>per group | Sample size per<br>group when<br>considering loss to<br>follow-up | Total sample<br>size of three<br>groups |
| --- | --- | --- | --- | --- |
| VSP vs Video training | 10.46 | 99 | 153 | 459 |
| VSP vs Manual training | 17.98 | 46 | 71 | 213 |

### 2.6. Study settings

We will conduct this study in Chongqing City, Sichuan Province, Guizhou Province, Yunnan Province, Jiangxi Province, Shanghai City, Guangdong Province, Qinghai Province, Hubei Province, Xinjiang Uygur Autonomous Region and other regions in China.

### 2.7. Randomization

A statistician will conduct randomization through the REDCap system, stratifying by region and randomly assigning study subjects recruited from each region to three intervention groups.

### 2.8. Variables

### 2.8. Blinding

This study employs a single-blind design. The participants are aware of the group to which they have been assigned but are unaware of the interventions of the other two groups. The assessors and data analysts are blinded to the group assignments, which helps to prevent subjective bias during assessment and data analysis.

## 3. Study Process, Data Collection and Management

The specific methods for data collection and management are as follows:

**Step 1: Informed consent/basic information/pre-intervention assessment**.

Study subjects will be invited to join a WeChat notification group specific to their region. The researcher will post links to the informed consent form, a basic information questionnaire, and an e-examination paper in the WeChat group, requesting participants to click the links, complete the informed consent form, and fill out the doctor’s basic information questionnaire along with the e-examination paper. The researcher then could obtain specific and extended competency scores based on the assessment results from the examination paper, while also completing the basic information of the medical organization.

**Step 2: Randomization and intervention**.

For those who complete the informed consent form and submit the baseline information, randomization will be performed. An independent statistician will generate the random sequence. The researchers or designated authorized personnel will randomly assign the participants from each region to one of the training groups: the VSP group, the case teaching video group, or the case teaching manual group, according to the random sequence and stratification by region. Each participant will receive a unique random enrollment number. The research assistant will create three different WeChat groups for different training methods for each province, asking all participants to complete the training within two weeks.

**Step 3: Collection of outcome indicators after intervention completion**.

(1) Within one week after the intervention, the investigators and study participants will determine the timing for collecting outcome indicators using Tencent Meeting, a video conferencing and online meeting platform widely used in China. The entire process will be audio and video recorded, with an estimated collection time of 30 minutes. After the investigators assess the participant using clinical vignette, a link will be sent to the Tencent Meeting chat box for the participant immediately. The link includes a questionnaire on disease diagnosis and treatment of the clinical vignette, an examination paper for specific and extended competency scores assessment, a team-developed patient-centered situational question test paper, and a questionnaire related to the implementation outcomes (Table 3). (3) Interviews will be conducted with doctors to explore the reasons for the effectiveness or lack thereof in each training group, involving 10 doctors per group for a total of 30. The interview questions include inquiries about the extent of participants’ engagement in the training, interest in the training type, perceived effectiveness, and reasons for these perceptions. For the VSP group, an additional question will be included: whether the VSP tool is perceived as too complicated or unfamiliar, leading to impatience. (4) Finally, the researcher will enter costs associated with developing and implementing the training into the REDCap system. All data will be captured and managed through REDCap.

**Table 3:**
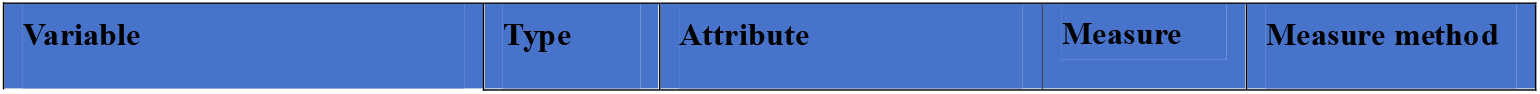
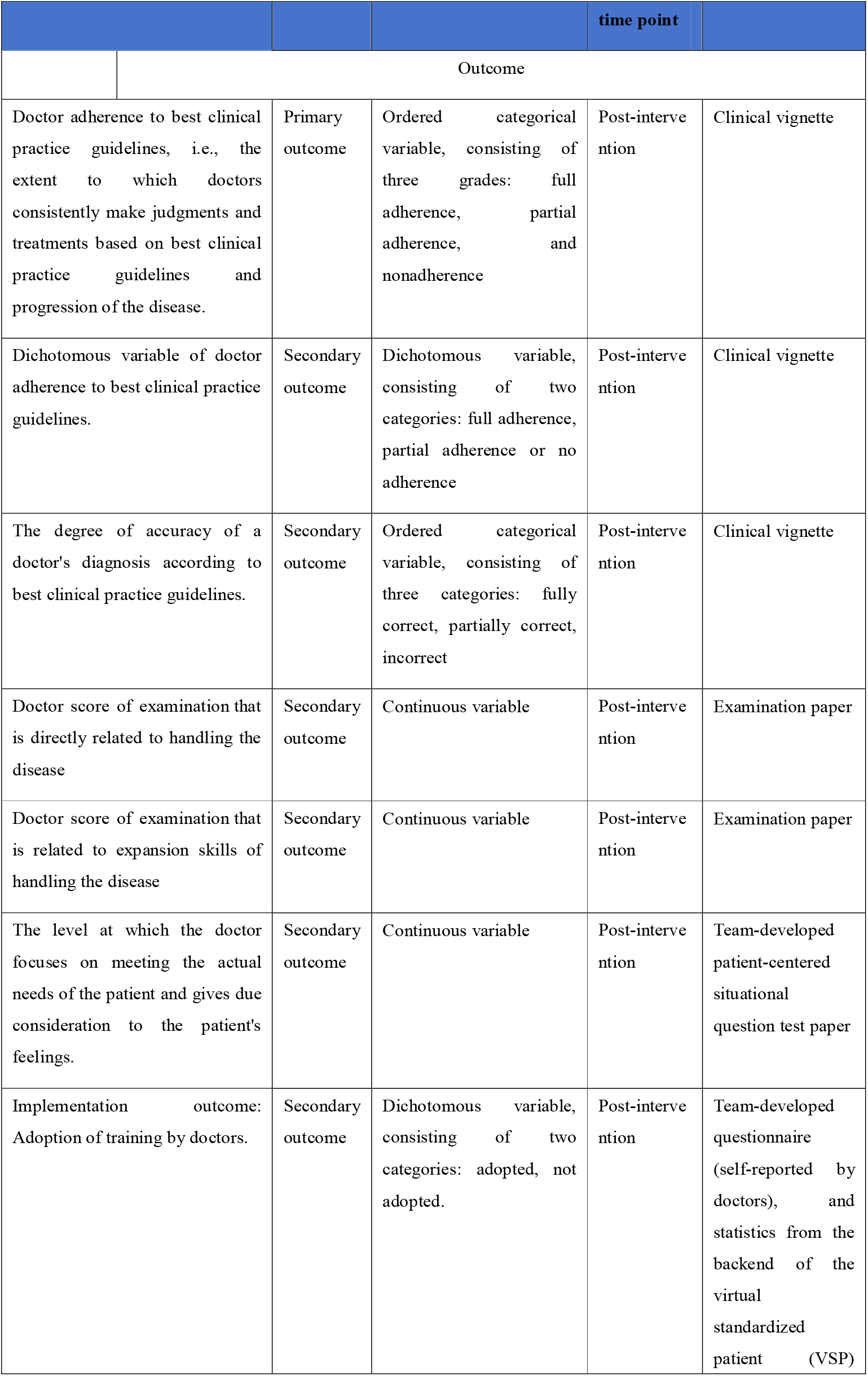
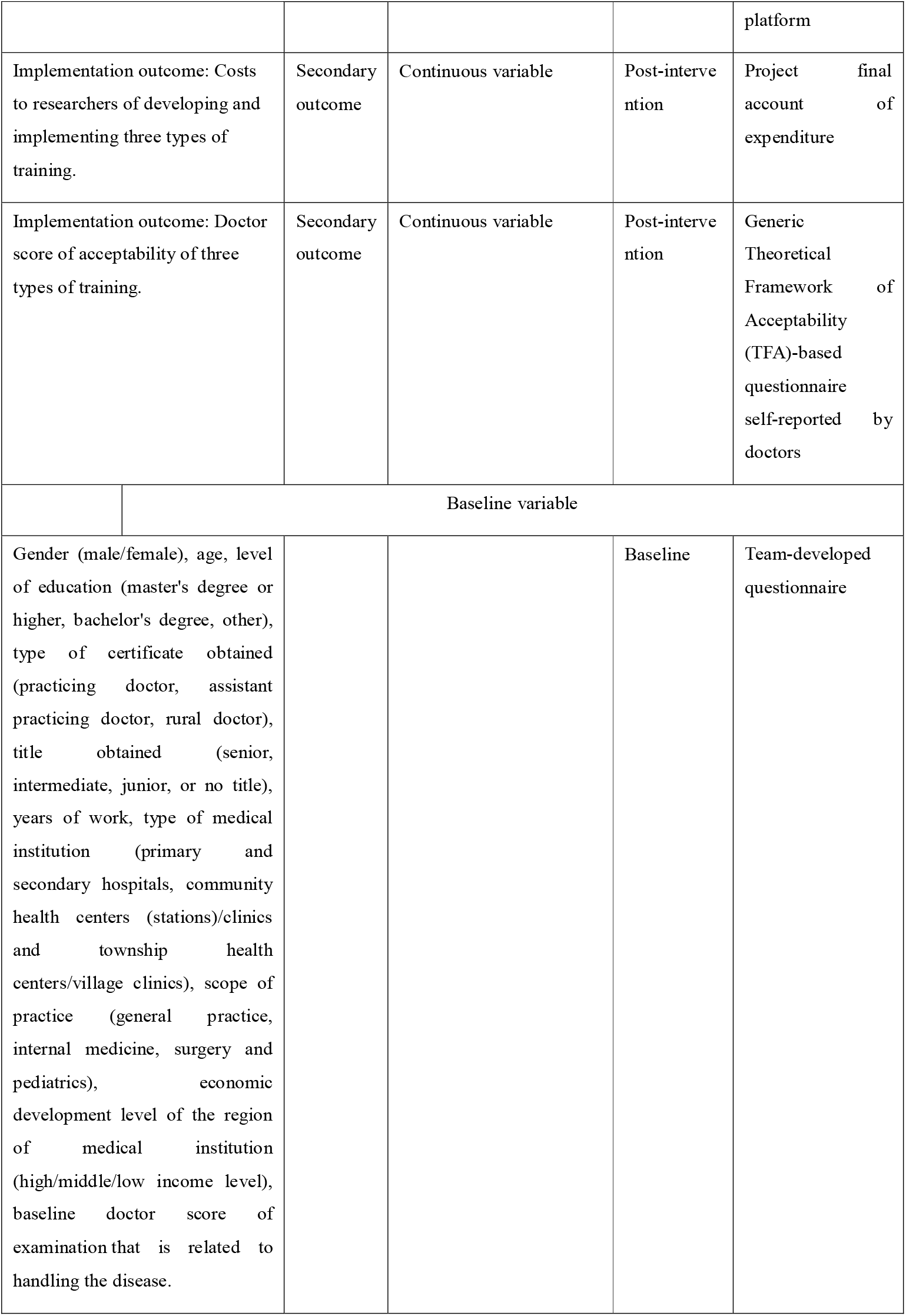
Variables information.

| Variable | Type | Attribute | Measure | Measure method |
| --- | --- | --- | --- | --- |

|  |  |  | time point |  |
| --- | --- | --- | --- | --- |
|  | Outcome |  |  |  |
| Doctor adherence to best clinical practice guidelines, i.e., the extent to which doctors consistently make judgments and treatments based on best clinical practice guidelines and progression of the disease. | Primary outcome | Ordered categorical variable, consisting of three grades: full adherence, partial adherence, and nonadherence | Post-intervention | Clinical vignette |
| Dichotomous variable of doctor adherence to best clinical practice guidelines. | Secondary outcome | Dichotomous variable, consisting of two categories: full adherence, partial adherence or no adherence | Post-intervention | Clinical vignette |
| The degree of accuracy of a doctor's diagnosis according to best clinical practice guidelines. | Secondary outcome | Ordered categorical variable, consisting of three categories: fully correct, partially correct, incorrect | Post-intervention | Clinical vignette |
| Doctor score of examination that is directly related to handling the disease | Secondary outcome | Continuous variable | Post-intervention | Examination paper |
| Doctor score of examination that is related to expansion skills of handling the disease | Secondary outcome | Continuous variable | Post-intervention | Examination paper |
| The level at which the doctor focuses on meeting the actual needs of the patient and gives due consideration to the patient's feelings. | Secondary outcome | Continuous variable | Post-intervention | Team-developed patient-centered situational question test paper |
| Implementation outcome: Adoption of training by doctors. | Secondary outcome | Dichotomous variable, consisting of two categories: adopted, not adopted. | Post-intervention | Team-developed questionnaire (self-reported by doctors), and statistics from the backend of the virtual standardized patient (VSP) |
|  |  |  |  | platform |
| Implementation outcome: Costs to researchers of developing and implementing three types of training. | Secondary outcome | Continuous variable | Post-intervention | Project final account of expenditure |
| Implementation outcome: Doctor score of acceptability of three types of training. | Secondary outcome | Continuous variable | Post-intervention | Generic Theoretical Framework of Acceptability (TFA)-based questionnaire self-reported by doctors |
| Baseline variable |  |  |  |  |
| Gender (male/female), age, level of education (master's degree or higher, bachelor's degree, other), type of certificate obtained (practicing doctor, assistant practicing doctor, rural doctor), title obtained (senior, intermediate, junior, or no title), years of work, type of medical institution (primary and secondary hospitals, community health centers (stations)/clinics and township health centers/village clinics), scope of practice (general practice, internal medicine, surgery and pediatrics), economic development level of the region of medical institution (high/middle/low income level), baseline doctor score of examination that is related to handling the disease. |  |  | Baseline | Team-developed questionnaire |

## 4. Data Analysis

### 4.1. Analysis of the Study Population

The analysis will consider two study populations. The intention-to-treat (ITT) population will be determined at the time of randomization. In the ITT analysis of this trial, participants will be tracked within their ITT subgroups. For example, if a participant studies materials from another group, they will still be analyzed according to their assigned group. The only exception will be late exclusions, such as those who do not meet the inclusion criteria, who will be excluded from the ITT analysis. The per-protocol (PP) population will be defined as participants who study the materials according to the protocol.

### 4.2. Statistical Analysis

Descriptive statistical analysis will be used for the basic information of the study participants and medical institutions. The primary outcome of the trial is the doctor adherence to best clinical practice guidelines, i.e., the extent to which doctors consistently make judgments and treatments based on best clinical practice guidelines and progression of the disease (an ordered categorical variable). The primary analysis will be based on the ITT population as defined above, using descriptive statistics of the participants number (n) and percentage (%).

Ordinal logistic regression with the treatment group as the study variable and the baseline variables as covariates will be used to estimate the odds ratio between the three groups and their corresponding two-sided 95% confidence intervals (95% CIs). The covariates include level of education (master’s degree or higher, bachelor’s degree, other), type of certificate obtained (practicing doctor, assistant practicing doctor, rural doctor), title obtained (senior, intermediate, junior, or no title), years of work, type of medical institution (primary and secondary hospitals, community health centers (stations)/clinics and township health centers/village clinics), baseline doctor score of examination that is related to handling the disease.

For sensitivity analysis, the missing values of the primary outcome of the three training groups will be imputed using multiple imputation method to verify the robustness of the primary analysis. Additionally, the subgroup analysis will be performed according to covariate category and median years of work. To address the issue of multiple comparisons, the Bonferroni correction will be applied to minimize the risk of type I errors.

For ordered categorical secondary outcomes, ordinal logistic regression will be employed, consistent with the approach used for the primary outcome. For dichotomous and continuous secondary outcomes, generalized linear models (GLMs) will be applied, with the treatment group as the study variable and the baseline variables as covariates. GLMs for dichotomous variables will use a binomial distribution with an identity link, estimating risk differences and their corresponding two-sided 95% CIs. GLMs for continuous variables will use a normal distribution with an identity link, estimating mean differences and their corresponding two-sided 95% CIs.

The economic evaluation of the three training schemes will be conducted using the incremental analysis method of cost-effectiveness analysis. Let CX represents the cost of training scheme X, CY represents the cost of training scheme Y, EX denotes the effectiveness of training scheme X, and EY denotes the effectiveness of training scheme Y, with the assumptions that CX>CY, EX>EY. The analysis process is as follows: (1) Rank all alternatives by cost from lowest to highest: CY, CX; (2) Identify the scheme with the lowest cost: Scheme Y is the scheme with the lowest cost; (3) Compare the higher-cost option with the lowest-cost option: △E = EX-EY, △ C = CX-CY. If △E/△C≥EY/CY, then scheme X is deemed cost-effective. This indicates that although training scheme X incurs a higher training cost compared to the control training scheme Y, it also yields a greater training effect. The additional cost of training scheme X is justified by the more substantial incremental benefits it provides. Therefore, training scheme X is considered cost-effective, potentially saving training costs for the entire population of doctors, and should be actively promoted and implemented.

## 5. Discussion

Artificial intelligence-driven virtual standardized patients (VSPs) use internet and virtual simulation technologies to replicate real clinical scenarios and symptoms via online platforms, including mobile applications and WeChat mini-programs. Doctors interact with VSPs to simulate clinical consultations, physical examinations, diagnoses, and treatment processes, specifically addressing gaps in primary pediatric care. VSPs offer flexibility, continuous assessment and feedback, risk-free practice, and standardized training, making them a promising tool for evaluating physician skills and supporting online medical education. This randomized controlled study will involve over 300 primary healthcare institutions across more than 10 provinces in China, encompassing both developed and underdeveloped regions, ensuring broad and representative participation. The study’s findings will provide valuable scientific evidence for the education and training of primary care doctors and inform strategies to enhance pediatric healthcare services.

## Data Availability

Not applicable.

## Abbreviations

VSPs: Virtual standardized patients
HFMD: Hand-foot-and-mouth disease
ITT: Intention-to-treat
PP: Per-protocol
CIs: Confidence intervals

## Acknowledgements

Not applicable.

## Authors’ contributions

YN M, HY L, YY C, Q Z, DR X and Y Z were involved in the study design. YN M, HY L and Y Z wrote the original draft of the protocol. HY L, WQ H and YC Z submitted the ethical review and registered the clinical trial. F F, Y L, X C, DR X and Y Z collaborated with the sites to develop the content of the intervention and the implementation plan. HY L, DL W and H H contributed to the sample size calculation and statistical analysis plan. XH W, DX Z and GB L contributed to the writing and critical review of the manuscript for important intellectual content. All authors read and approved the final manuscript.

## Availability of data and materials

Not applicable.

## Declarations

### Ethics approval and consent to participate

This study will be performed strictly in accordance with the Declaration of Helsinki and follows the principles of informed consent and voluntary participation. This study has been approved by the Biomedical Ethics Committee, Southern Medical University (NFYKDX003). Written informed consent will be obtained from each participant before data collection.

## Consent for publication

Not applicable.

## References

1. Xu BX, Lin XD, Yao WG. Children’s intention to seek healthcare in primary healthcare settings and associated determinants: an analysis using the Anderson’s behavioral model of health services use. Chinese General Practice. 2022;25(22):2766–2772.

2. Wang SC, Chen YF, Gu H, et al. The concept, models, and cases of virtual patients in clinical trial simulation. Chinese Journal of New Drugs. 2024;33(09):911–916.

3. Wen ZH. Diagnosis and prevention strategies for pneumococcal pneumonia in children. Chinese Journal of Clinical New Medicine. 2021;14(03):238–244.

4. Xu H, Chen M, Sun YF, et al. Pathogen and drug sensitivity analysis of bronchoalveolar lavage fluid in children with severe community-acquired pneumonia in Guiyang. Guangdong Medical Journal. 2020;41(23):2394–2397.

5. Jin Y, Mankadi PM, Rigotti JI, et al. Cause-specific child mortality performance and contributions to all-cause child mortality, and number of child lives saved during the Millennium Development Goals era: a country-level analysis. Glob Health Action. 2018;11(1):1546095.

6. Hao JY, Zhu D, Ma X, et al. Analysis of the causes and clinical characteristics of persistent and chronic diarrhea in children. Chinese Journal of Pediatric Emergency Medicine. 2023;30(12):937–941.

7. Zhang J, Li XH, Li L, Shang XF, et al. Etiology and epidemiology of hand, foot and mouth disease in China. Chinese Journal of Epidemiology. 2022;43(5):771–783.

8. Liu YQ, Tong WB, Ma XZ, et al. Epidemiological and pathogenic characterization of hand, foot, and mouth disease in Sichuan Province from 2017 to 2021. Modern Preventive Medicine. 2024;51(10):1742–1747.

9. Chen CY, Qiu LX, Zhao ZY, et al. Economic burden and influencing factors of hand, foot, and mouth disease in Bao’an District, Shenzhen. Nursing Research. 2024;38(07):1262–1268.

10. You H, Cao T, Sheng Y. Analysis of compliance behavior and influencing factors in children undergoing laparoscopic surgery for acute appendicitis. Chinese Journal of Social Medicine. 2024;41(03):312–315.

11. Zhang N, Jiang X. Clinical progress and treatment advances in pediatric acute appendicitis. Journal of Clinical Pediatric Surgery. 2020;19(11):1042–1046.

12. Liu DZ, Li JY, Geng H. Research progress in the diagnosis, treatment, and postoperative enhanced recovery of pediatric acute appendicitis. Smart Healthcare. 2024;10(9):29–32.

13. Yang M, Qian SY. Clinical manifestations and diagnosis of respiratory failure. Practical Journal of Pediatric Clinical Medicine. 2010;25(4):234–236.

